# Implementation of COVID-19 Preventive Measures in Primary and Secondary Schools Following Reopening of Schools in Autumn 2020; A Cross-Sectional Study of Parents’ and Teachers’ Experiences in England

**DOI:** 10.1101/2021.06.03.21258289

**Authors:** Zahin Amin-Chowdhury, Marta Bertran, Meaghan Kall, Georgina Ireland, Felicity Aiano, Annabel Powell, Samuel E Jones, Andrew Brent, Bernadette Brent, Frances Baawuah, Ifeanychukwu Okike, Joanne Beckmann, Joanna Garstang, Shazaad Ahmed, Neisha Sundaram, Chris Bonell, Sinead Langan, James Hargreaves, Shamez N Ladhani

**Affiliations:** Immunisation and Countermeasures Division, Public Health England, Colindale, London NW9 5EQ, UK; National COVID-19 Epidemiology Cell, Public Health England, London, UK; Nuffield Department of Medicine, University of Oxford, Oxford University Hospitals NHS Foundation Trust, Oxford, UK; Oxford University Hospitals NHS Foundation Trust, Oxford, UK; Derbyshire Children’s Hospital, University Hospitals of Derby and Burton NHS foundation Trust, Derby, UK; Specialist Children & Young People’s Services, East London NHS Foundation Trust, London, UK; Birmingham Community Healthcare NHS Trust & University of Birmingham, Birmingham, UK; Department of Virology, Manchester Foundation Trust, Manchester, UK; Department of Global Health and Development, London School of Hygiene & Tropical Medicine, London, UK; Department of Non-communicable Disease Epidemiology, London School of Hygiene & Tropical Medicine, London, UK; Department of Public Health, Environments and Society, London School of Hygiene & Tropical Medicine, London, UK; London School of Hygiene & Tropical Medicine, London, UK; Paediatric Infectious Diseases Research Group (PIDRG), St. George’s University of London, Cranmer Terrace, London SW17 0RE, UK

**Keywords:** preventive measures, schools, COVID-19, SARS-CoV-2

## Abstract

**Objective:** The main objective was to assess implementation of and ease of implementation of control measures in schools as reported by staff and parents.

**Design:** Cross-sectional study.

**Setting:** Staff and parents/guardian participants in the 132 primary schools and 20 secondary schools participating in sKIDs and sKIDsPLUS surveillances.

**Main outcome measure:** Prevalence of control measures implemented in Autumn 2020, parental and staff perception of ease of implementation and acceptability of conducting school surveillance studies.

**Results:** In total, 56/152 (37%) schools participating in Public Health England’s sKIDs study of COVID in schools accepted the invitation to participate in the survey. By 28 December 2020, 1,953 parent and 986 staff respondents had completed the online questionnaire. While more than half the parents were positive about their children returning to school, roughly a third reported being a little anxious. 90% and 82% of primary and secondary school parents were either completely or partly reassured by the preventive measures implemented in their schools. Among staff, 80% of primary staff and 87% of secondary school staff felt that they were at higher risk of COVID-19 because of their profession; only 52% of primary school staff and 38% of secondary school staff reportedly felt safe. According to the teaching staff, most preventive measures were well-implemented apart from requiring 2-metre distancing between staff. For students, maintaining the 2-metre distance was reported to be particularly difficult. By extension, secondary schools also struggled to maintain small groups at all times or ensuring that the same staff were assigned to each student group (a problem also commonly reported by parents).

**Conclusions:** Variable implementation of infection control measures was reported by staff and parents. Whilst the majority were not worried about returning to school, some parents and staff, were concerned about returning to school and the risks posed to children, staff and household members.

**Strengths and limitations of this study:** *Strengths:*

- This study is one of the few to investigate school staff and parents’ perceptions of the implementation of control measures implemented following the reopening of schools in England.
- The early establishment of COVID-19 surveillance in primary and secondary schools in the summer term 2020 provided a cohort to rapidly evaluate the experiences of parents and school staff during the autumn term before schools were required to close for the subsequent national lockdown.

*Limitations:*

- As the questionnaire and information provided was available in English only, there is likely to be an under-representation of families for whom English was not their main language.
- Some school responses were only provided by one participant so may not necessarily be representative of the whole school.
- Although the surveillance included schools recruited nationally, a convenience sample was used and as such may not be representative of all primary and secondary schools in England.

## Background

Children have been, comparatively less affected by coronavirus disease 2019 (COVID-19) caused by severe acute respiratory syndrome coronavirus 2 (SARS-CoV-2), representing only 1-3% of confirmed COVID-19 cases, with very few hospitalisations and deaths.^2-4^ Whether or not schools should remain open or closed remains a contentious topic for debate in the COVID-19 pandemic.^5^

Early in the pandemic, the role of children in transmission of SARS-CoV-2 was still unclear and many countries implemented national lockdowns which included school closures.^6^ In the UK, children of key workers and vulnerable children, however, continued to attend school throughout the first lockdown. Subsequently, from 01 June 2020, some school years (nursery, reception, year 1 and year 6) returned to school, followed by some secondary school years (years 10 and 12) from 15 June 2020, although school attendance was not mandatory.^7^ The second half of the summer term was continued until 18 July 2020 before summer holidays began. During this period, strict physical distancing and infection control measures were implemented in schools, including limiting class sizes to small numbers, who remained in strict social bubbles that did not interact physically or social with other bubbles in school.^8^ The success of the summer half-term (where few cases and outbreaks of COVID-19 were reported) contributed to the wider re-opening of all schools with full attendance in the autumn term, which started in September 2020.^9^

The large number of students attending primary and secondary schools during the autumn term was likely to raise significant challenges for implementing and reinforcing physical distancing and infection control measures. In addition to the challenges of maintaining infection control measures in educational settings, community SARS-CoV-2 infection rates were higher at the start of the autumn term compared to the previous summer half-term.^10^ This in turn raised concerns about increased risk of SARS-CoV-2 introduction into education settings, via outbreaks that result in isolation of large class bubbles or potential closures if infection could not be controlled through current national guidelines and recommendations.^11^

To better understand the impact of SARS-CoV-2 in educational settings, Public Health England (PHE) has been conducting SARS-CoV-2 surveillance since the start of the pandemic in England which has included swabbing and serological sampling in selected primary and secondary schools.^9^ As part of this surveillance, we assessed the experiences and challenges of returning to school during the Autumn term by inviting the schools taking part in PHE school studies to participate in an online survey aimed at teaching staff and parents two months after the students returned to school in September 2020.

## Methods

### Schools surveillance

As part of national surveillance, PHE initiated enhanced surveillance in 132 primary schools which were selected as previously described^12^ in five sites across England (East London, North and West London, Derby, Oxford and Manchester), during the summer half-term where staff and students were tested for SARS-CoV2 infection through weekly swabbing or blood sampling for SARS-CoV-2 antibodies at the beginning and end of the summer half-term (sKIDs studies). From September 2020, surveillance was extended to include 20 secondary schools (sKIDsPLUS study).^13^

### Study design

A cross-sectional survey was conducted among staff – including teachers, teaching assistants and senior leadership teams – and parents/guardians in the 132 primary schools and 20 secondary schools participating in sKIDs and sKIDsPLUS surveillance. PHE’s studies. These schools were invited to take part in the online survey during the first week of November 2020, when schools returned from the half-term holidays. Schools expressing an interest in taking part were provided with an online link to forward to the teaching staff and all the parents in that school, irrespective of whether they were taking part in the sKIDs surveillance. The surveys were disseminated on 10 November 2020 and up to 5 email reminders were sent to encourage participants to complete the questionnaire until the last day of the survey on 28 December 2020.

### Questionnaire design

Staff and parent questionnaires were adapted from those used during summer 2020 in a survey for headteachers and designed using Snap Professional 11 (SnapSurvey).^14^ Participants were provided with a list of preventive measures and were asked whether any of the measures were being implemented at their school as far as they were aware.

Interventions were grouped into those related to students, staff or the classroom and school environment. Parents were asked to rate to what extent their child was able to follow these preventive measures. Similarly, staff were asked the ease of implementation of the control measures. The sKIDs and sKIDsPLUS studies were approved by PHE’s ethics committee as a part of its responsibility to investigate SARS-CoV-2 infections among children in educational settings. Participants were informed that the survey was anonymised and voluntary, completion and return of questionnaire was taken as implied consent.

### Data analysis

Questionnaire responses from SnapSurvey were imported into Stata 15.1 (StataCorp, Tx). Data were cleaned and a descriptive analysis performed, stratified by parent/staff category and primary/secondary school. School profiles obtained from Department for Education data were used to compare school demographic characteristics including type of school, school size, percentage of students on free school meals and percentage of persistent absence in responding schools and non-responding schools.^15^ To report questions related to control measures at a school level, responses were weighted such that for schools with multiple responses, the contribution of each response summed to 1 in the weighting to compensate for overrepresentation of schools with more responses than others. Categorical variables are presented as proportions and compared using chi-squared or Fisher’s Exact tests, where appropriate. Data that did not follow a normal distribution are described as median with interquartile ranges and compared using the Mann Whitney *U* test.

## Results

### Characteristics of parent and staff respondents

In total, 152 schools participating in sKIDs and sKIDsPLUS across England were contacted, of which 56 (39%) participated in the survey. By 28 December 2020, 1,953 parent and 986 staff respondents had completed the online questionnaire. Parents from 41/132 (31%) primary schools with a mean response of 24 (range 1-109) and 15/19 (79%) secondary schools (mean: 64; range (2-173) responded to the parent/guardian survey and staff from 56/132 (42%) primary schools (mean: 8; range (1-59) and 17/19 (90%) secondary schools (mean: 30; range (10-66) responded to the staff survey. Response rates were higher in secondary schools which were larger but otherwise characteristics were broadly similar between responding and non-responding schools (**Supplementary Table 1**). Most primary school teachers (81%) reported working in bubble sizes of 26-30 (45%) or >30 (37%), while almost two-thirds of secondary school teachers (63%) reported working in bubble sizes >30 or “other”, which constituted the whole year group in most cases (data not shown).

#### Parents

Of those parents/guardians that responded, 999 (51%) reported that their child attended primary school and 954 (49%) attended secondary school. Approximately half of all respondents (1,029/1,953) reported that either they or their child’s other parent/guardian was a key worker. Over a third of primary school parents (345/999, 35%) reported their child was eligible to receive free school meals, compared to only 13% (120/954) of secondary school parents (**Supplementary Table 2**). Only 45/1,953 (2%) parents overall (6/999 (0.6%) in primary and 39/954 (4%) in secondary) reported their child had tested positive for SARS-CoV-2 infection (data not shown).

#### Staff

In total, there were 471 (48%) respondents from primary schools and 515 (52%) respondents from secondary schools (**Supplementary Table 2**). Over half the primary school respondents were teachers (268, 57%) compared to 341 (66%) of secondary school respondents. Conversely, there was a higher proportion of teaching assistants in primary (95, 19%) compared with secondary (147, 31%) in secondary schools (**Supplementary Table 2**). Senior leadership team respondents including headteachers, deputy and assistant headteachers comprised 56 (12%) primary and 51 (10%) secondary staff respondents.

### Implementation of preventive measures at school

#### Student measures according to parents

Parents from 93% (51/56) reported that regular hand cleaning for students was the most frequently reported measure (70, 94%), and more frequently reported in primary (39/41, 96%) than in secondary schools (13/15, 84%) (**Table 1**). Other student measures that more frequently reported by primary than secondary school parents included respiratory hygiene (30/41, 73% vs 13/15, 63%), keeping students within the same small groups at all times (28/41, 69% vs 8/15 55%) and ensuring the same teacher/staff member was assigned to each student group (29/41, 71% vs 6/15, 38%). Measures less commonly implemented in primary and secondary schools included daily/weekly rota for attending school was (11/41, 26% and 6/15, 37%, respectively) and were daily temperature checks (2/41, 4% and 2/15, 10%, respectively). In line with guidance, over three-quarters (12/15, 78%) of secondary parents reported that their child was required to wear a face covering while at school, compared to only 2/41 (4%) in primary (**Table 1**).

**Table 1.** Student measures implemented as reported by parent/staff, respectively

|  | Parent |  | Staff |  |
| --- | --- | --- | --- | --- |
|  | Primary school<br>n (%)<br>N=41 | Secondary school<br>n (%)<br>N=15 | Primary school<br>n (%)<br>N=56 | Secondary school<br>n (%)<br>N=17 |
| **Requiring maintenance of 2m distance from others for students** | 12 (30.3%) | 8 (51.4%) | 13 (23.8%) | 6 (32.8%) |
| **Requiring regular hand cleaning for students** | 39 (96.0%) | 13 (84.4%) | 55 (98.8%) | 15 (89.2%) |
| **Ensuring students catch cough or sneezes with tissue or arm** | 30 (73.1%) | 9 (63.2%) | 52 (92.3%) | 11 (65.5%) |
| **Keeping students with the same small groups at all times each day** | 28 (68.8%) | 8 (55.0%) | 43 (76.6%) | 9 (54.5%) |
| **Ensuring that the same teacher(s) and other staff are assigned to each student group** | 29 (71.2%) | 6 (37.6%) | 37 (66.5%) | 3 (14.9%) |
| **Students attending school on a daily or weekly rota** | 11 (26.2%) | 6 (36.7%) | 8 (15.1%) | 3 (16.8%) |
| **Students required to wear face masks or face coverings while at school** | 4 (8.5%) | 12 (78.4%) | 2 (3.7%) | 16 (92.1%) |
| **Daily temperature checks for students** | 2 (4.4%) | 2 (10.0%) | 4 (6.6%) | 1 (7.7%) |
| **Ensuring students who have coronavirus symptoms, or have someone at home who does, stay home** | - | - | 54 (97.1%) | 16 (96.1%) |

#### Parents’ perception of their child’s compliance of preventive measures

Among parents of primary school children, regular hand washing (24/41, 58%) and respiratory hygiene (13/41, 33%) were reported to be easiest to follow all the time (**Figure 1**). While wearing a face mask was not recommended in the guidelines for children under the age of 11 years, 25% (10/41) of parents reported their child wore one on public transport always or most of the time. Compared to primary schools, parents of secondary school children reported higher compliance of all preventive measures in their children. Among parents of secondary school children, most reported that their child always wore a face mask/covering in public areas (10/15, 73%) or when on public transport (9/15, 60%). The most difficult measure to follow all the time among children of both primary (4/15, 30%) and secondary schools (7/14, 19%) was keeping a 2-metre distance from others when outside of the home, although most parents of primary (22/41; 53%) and secondary school (11/15, 72%) children reported that their child kept a 2-metre distance at least most of the time (**Figure 2**).

**Figure 1.** Child’s compliance of preventive measures as reported by parents

**Figure 2.** Perceived ease of implementation of staff measures, by staff

#### Student measures according to staff

Almost all staff in primary (54/56, 97%) and secondary schools (16/17, 96%) reported that the school ensured students with COVID-19 symptoms would be required to stay at home (**Table 1**). Similarly, regular hand washing was also reported to be widely implemented (55/56, 99% and 15/17, 89%, respectively). Some measures were rarely reported to be implemented, such as students attending school on daily/weekly rota (8/56, 15% and 3/17, 17%, respectively) and daily temperature checks for students (4/56, 7% and 1/17, 8%, respectively). Other measures reported at very different frequencies in primary and secondary schools included wearing face masks/coverings by students (16/17, 92% and 2/56, 4%, respectively) as per national guidance, and ensuring the same staff are assigned to each student group (37/56, 67% and 3/17, 17%, respectively).

#### Staff perception of students’ compliance of preventive measures

When asked how challenging staff found the implementation of student measures, the most challenging measure was requiring students to maintain 2-metre distancing in both primary (37%) and secondary (58%) schools. Where implemented, the easiest measures reported by staff were daily temperature checks for students (71% and 88%, respectively) and students attending school on a weekly/daily rota (70% and 52%, respectively). Regular hand cleaning was reported by staff to be easier to implement in primary (45%) than in secondary (34%) schools (data not presented.

#### Preventive measures for staff

The vast majority of primary schools (54/56, 97%) and secondary schools (15/17, 93%) as reported by staff had received guidance by the school on what to do if a student or staff has COVID-19 symptoms. The majority of staff reported that their school required regular hand cleaning for staff (97% in primary/secondary) and requiring a 2-metre distance from others (83%) (**Table 2**). Approximately half of primary (29/56, 51%) and secondary (8/17, 47%) reported that they had stopped all in-person staff meetings. Facemasks/coverings for staff members were reported to be implemented in 92% (16/17) secondary schools and 38% (22/56) primary schools (42%). Other measures not commonly implemented by either primary or secondary schools included staff being advised to work from home if their job could be done from home (7-13%) or if they lived in a household with an extremely clinically vulnerable individual (13-15%). Less than half the primary (22/46, 39%) and secondary (7/17, 41%) school staff reported that the school advised not to attend work or to work from home if they themselves were clinically vulnerable (**Table 2**).

**Table 2.**
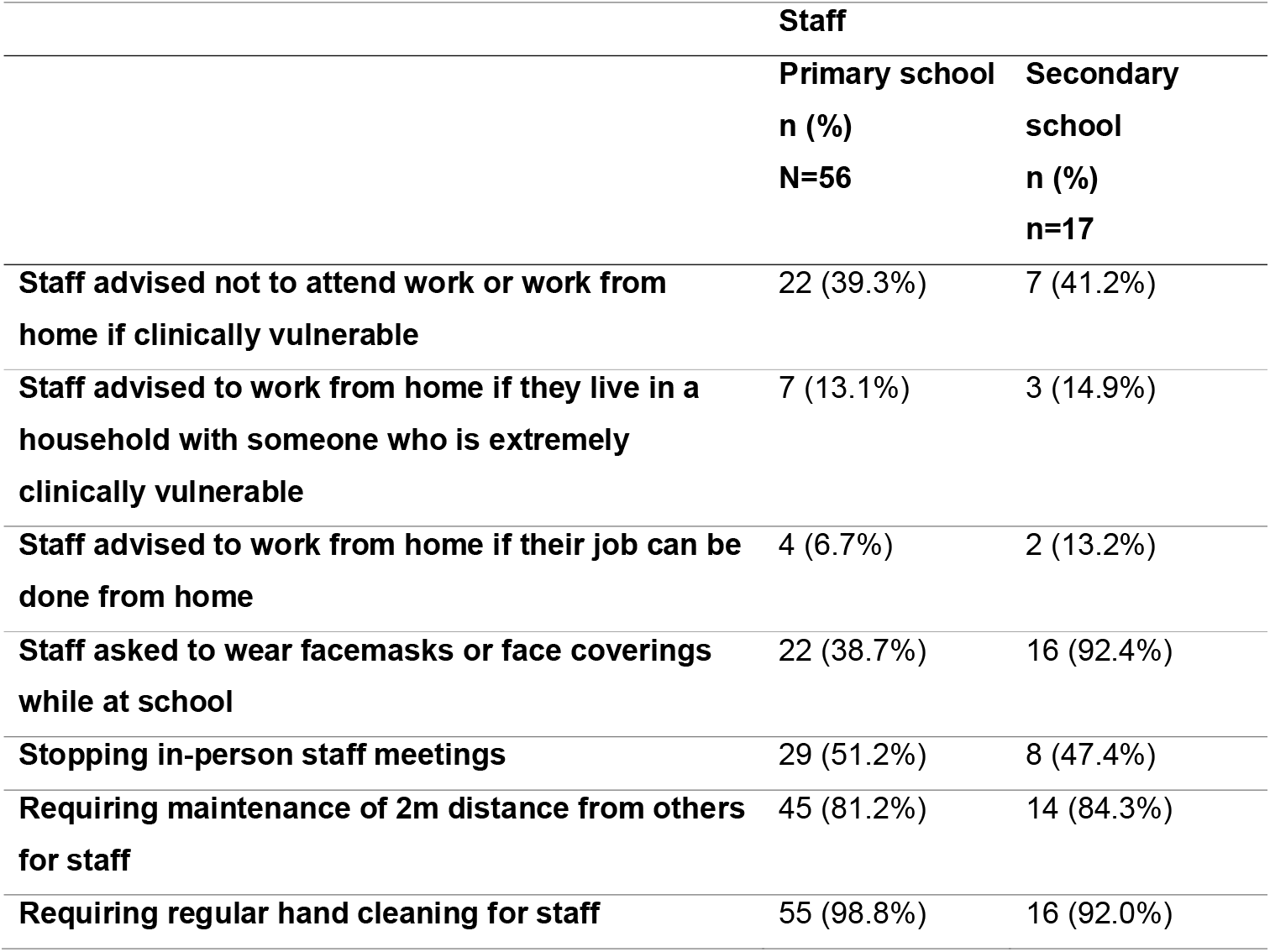
Preventive measures implemented at school for staff

Most (16/17, 92%) of secondary schools reported that they were advised to wear a face mask/covering outside classrooms, of which, 17% reported that they were required to wear them all the time. In comparison, 33% primary school staff reported being required to wear a facemask/covering outside classrooms and only 6% reported to being required to wear them all the time. Of the remaining, 25% reported that they were given the option to wear a facemask/covering or not.

When asked how challenging it was to implement preventive measures for staff, the 2-metre distancing was the most challenging, with 27% of primary and 36% of secondary school staff reporting that it was ‘very challenging’ (**Figure 2**). In contrast, regular hand cleaning was the easiest to implement (79% and 77%, respectively). Most primary and secondary school staff reported that there were ‘some challenges’ to staff working from home if clinically vulnerable (49%), if they were living with someone clinically vulnerable (55% vs 58%) or if their work could be done from home (49% vs 61%).

### School and the environment measures

#### Staff reporting of preventive measures in the school and classroom

Fitting hand sanitisers at the school entrance, stopping large gatherings and staggering break times for different classes were among the most commonly reported measures by staff of both primary and secondary schools (>85%). Some measures were more commonly reported to be more challenging by primary than secondary school staff, such as requiring 2-metre distancing for parents dropping off or picking up children (87% vs 29%, respectively) and staggering drop-off or collection times (92% vs 72%, respectively). Other measures were not as commonly reported by staff, such as removing/disabling air flow hand driers from toilets (27-33%) (**Table 3**).

**Table 3.** Preventive measures implemented at child’s school (classroom and school environment)

|  | **Primary**<br>**school**<br>**n (%)**<br>**N=41** | **Secondary**<br>**school**<br>**n (%)**<br>**N=15** | **Primary**<br>**school**<br>**n (%)**<br>**N=56** | **Secondary**<br>**school**<br>**n (%)**<br>**N=17** |
| --- | --- | --- | --- | --- |
| **Classroom measures** |  |  |  |  |
| **Fitting hand sanitisers in classrooms** | 19 (47.4%) | 9 (60.7%) | 40 (72.2%) | 13 (76.4%) |
| **Maintaining space between seats and desks** | 22 (52.9%) | 7 (47.6%) | 28 (50.8%) | 7 (43.2%) |
| **Removing non-essential objects from classrooms** | - | - | 35 (62.0%) | 7 (41.2%) |
| **Removing soft furnishings and toys that are hard to clean** | - | - | 39 (69.0%) | 4 (26.3%) |
| **Cleaning frequently touched surfaces** | - | - | 50 (89.2%) | 15 (89.2%) |
| **Scheduling more lessons and activities outdoors** | 13 (31.3%) | 2 (11.9%) | 14 (25.4%) | 1 (3.4%) |
| **Ensuring students use the same classroom throughout the day** | - | - | 49 (87.7%) | 7 (39.1%) |
| **Ensuring students do not share equipment/learning materials in classrooms** | - | - | 36 (65.1%) | 12 (70.4%) |
| **Ensuring students do not carry equipment/learning materials between home and school** | 19 (46.0%) | 3 (20.0%) | 33 (58.3%) | 2 (13.7%) |
| **School and the environment measures** |  |  |  |  |
| **Introducing one-way systems in school corridors** | 19 (46.3%) | 9 (63.1%) | 31 (55.7%) | 11 (63.8%) |
| **Fitting hand sanitisers at the school entrance** | 17 (42.5%) | 8 (56.6%) | 50 (89.3%) | 15 (88.6%) |
| **Removing / disabling air flow hand driers from toilets** | - | - | 20 (36.5%) | 4 (23.4%) |
| **Staggering drop-off and collection times** | 31 (74.7%) | 9 (60.6%) | 50 (89.0%) | 12 (70.6%) |
| **Staggering break times for different classes** | - | - | 51 (91.1%) | 14 (85.2%) |
| **Stopping large gatherings of students e.g. assemblies** | 31 (76.0%) | 10 (68.8%) | 53 (93.8%) | 15 (88.5%) |
| **Stopping team sports** | 14 (35.4%) | 6 (42.1%) | 30 (53.9%) | 8 (45.0%) |
| **Requiring 2m distancing for parents dropping off or picking up children** | 28 (67.3%) | 4 (27.9%) | 49 (88.3%) | 4 (23.4%) |
| **Allowing only one parent to accompany child to school** | 22 (53.7%) | 3 (19.1%) | - | - |
| **Encouraging parents and children to not travel to school using public transport** | 9 (21.5%) | 5 (30.2%) | - | - |

The majority of staff in primary and secondary schools reported that hand sanitisers were fitted in their classroom (40/56, 72% and 13/17, 76%, respectively) and that touch surfaces were frequently cleaned (50/56, 89% and 15/17, 89%, respectively) (**Table 3**). Other measures were more frequently reported by primary school staff than secondary school staff, such as removing soft furnishings and toys that are hard to clean (39/56, 69% vs 4/17, 31%, respectively), ensuring students were in the same classroom all day (49/56, 87% vs 7/17, 39%, respectively) and scheduling more lessons and activities outdoors (14/56, 25% vs 1/17, 3%, respectively) (**Table 3**).

Fitting hand sanitisers in the classrooms, removing/disabling air flow hand driers from toilets and removing soft furnishing/toys that are hard to clean were some of the easiest measures to implement in both primary and secondary schools (>70%). Maintaining space between seats and desks was reported by more than half the primary (52%) and secondary school staff (60%) to have ‘some challenges’. Among primary school staff, the 2-metre distancing at drop-off/collection was reported to be the most challenging with the majority reporting that this measure had ‘challenges’ (52%) or was ‘very challenging’ (31%). Among secondary school staff, introducing a one-way system in corridors and staggering break times were reported to be most challenging with more than half reporting ‘some challenges’ (55%) (data no presented.

## Discussion

### Summary of key findings

The early establishment of COVID-19 surveillance in primary and secondary schools provided a unique opportunity to rapidly evaluate the experiences of parents and teaching staff following the full reopening of all schools in England in September 2020. More than 150 schools taking part in sKIDs across England were invited to take part in the questionnaire survey and 58 agreed to forward the online questionnaire links to their staff and parents of students attending their schools. Reassuringly, more than half of parents were positive about their children returning to school, similar to findings of a similar US survey of parents.^1^ However, around a third of parents reported being a little anxious, while 13% and 16% of primary and secondary school parents, respectively, reported being extremely anxious about their children returning to school. In general, when asked about preventive measures implemented in their schools, parents reported variable rates of implementation for their schools. In primary schools, staggering drop-off and collection times and stopping large gatherings of students such as assemblies were the only preventive measures reported by more than 75% of parents, and the latter was the only measure reaching this frequency in secondary schools. Overall, however, 90% and 82% of primary and secondary school parents were either completely or partly reassured by the preventive measures implemented in their schools.

Among staff, a significant finding of this survey was that 80% of primary staff and 87% of secondary school staff felt that they were at higher risk of COVID-19 because of their profession. Indeed, only 52% of primary school staff and 38% of secondary school staff felt safe at school despite the implementation of a wide range of social distancing and infection control measures. According to the teaching staff, preventive measures for staff were variably implemented, apart from regularly hand cleaning, maintaining a 2-metre distance between staff members and, for secondary school staff, wearing facemasks or face coverings while at school. In particular, most staff did not feel like they were given the option to work from home if possible, even if there was a clinical reason to do so. According to the teaching staff, most preventive measures were well-implemented apart from requiring 2-metre distancing between staff.

For preventive measures involving students, too, maintaining the 2m distance was found to be particularly difficult to implement for both primary and secondary schools, while secondary schools also struggled to maintain small groups at all times or ensuring that the same staff were assigned to each student group – a problem also commonly reported by parents. This was also reflected in teaching staff experiencing difficulties with maintaining space between seats and desks in both primary and secondary schools. Another problem faced particularly by secondary school teachers was ensuring that the students used the same classroom throughout the day and ensuring that the students do not carry materials between home and school. Other measures were implemented to a variable extent, except for parents dropping off or picking up secondary school students, which may be because most secondary school students are not picked up by their parents.

### Comparison with published literature

It is difficult to compare the current experience of educational staff and parents of primary and secondary school students with other countries, each with different community SARS-CoV-2 infection rates and implementation of local infection control measures to mitigate the risk of infection in their settings. We also used a standardised questionnaire to rapidly collect qualitative information from a large number of staff and students in school that were already participating in SARS-CoV-2 surveillance, which contrasts with other studies which mainly involved detailed interviews with a small number of staff and parents.

One cross-sectional survey study in England, using a convenience sample of 442 participants, measured parental perceptions of control measures implemented in their child’s school in June. Their findings suggested that suboptimal practices were widespread, with only half of parents reporting hand-washing or hand gel dispensing facilities at school entrances and in classrooms and almost 40% reporting class sizes being larger than the recommended fifteen.^16^ With the full reopening of schools in the autumn term, limitation of class sizes into small distinct bubbles was no longer possible, especially for secondary schools, where classes are much larger than primary school classes.

In England, the experiences of the current autumn term when all the students returned to school is very different to the previous summer half-term, when only some primary and secondary school years, returned to school and with small class bubble sizes. At that time, detailed interviews with headteachers of the sKIDs schools identified different challenges in implementing infection control measures, including difficulties in prioritising teaching because of the additional requirement and practices, physical space constraints, staffing issues, finances, lack of adequate protective equipment and parent. The inability to maintain the 2-metre distance between the students and between students and staff, especially in primary schools, has been a consistent finding and not only challenging to implement but also considered incompatible with good teaching, especially in early-years classrooms.^17^

The autumn term is different to the previous summer half-term for a number of reasons. The number of students returning to school was much higher, increasing the challenges already encountered in maintaining physical distancing and infection control measures. At the same time, community infection rates were much higher between September and December 2020 than they were in June 2020, with increased numbers of cases in school-aged children,^18^ and outbreaks in primary and secondary schools.^19^ This had a large impact on the number of staff and student contacts required to self-isolate as part of the contact bubbles. Often whole classes and year groups had to self-isolate following a single confirmed case, and many staff and students had to self-isolate multiple times because they were contacts of different cases in their bubble. This was disruptive not only for the self-isolating students but also the remaining students because of the inconsistencies in school attendance and teaching staff.

### Strengths and limitations of this study

The strength of this survey was the establishment of good relationships with primary and secondary school taking part in school surveillance studies which enabled a good response and the timely implementation of the survey during the autumn term before schools were required to close for the subsequent national lockdown. However, there are some limitations. We did not assess responses by demographics, such as ethnicity, or socio-economic status, which are very likely to influence questionnaire responses among staff and students, as has been reported elsewhere.^1^ Instead, we provided a summary of the demographics and results for all participants combined to ensure that the key messages reflect the group as a whole. Additionally, not all schools agreed to forward the questionnaire to their staff and parents, mainly because they too busy but this may have also introduced selection bias if those that felt less prepared were less willing to participate in the questionnaire survey, perhaps anxious about what this might reveal. While participating primary schools had a broad geographical spread across England, they are not representative of all primary and secondary schools in England. Primary schools participating in sKIDs were selected because they re-opened with at least 30 students in attendance during the summer half-term. Similarly, secondary schools were identified for sKIDsPLUS because they were located in five regions where paediatric teams were available for taking blood samples for antibody testing from staff and students.

Of those who took part, it is likely that there was an under-representation of families for whom English was not their main language as well as those with limited access to digital platforms and the internet. Nonetheless, parents from 56 schools and staff from 73 schools agreed to take part and nearly 3,000 questionnaires were completed within 3 weeks, allowing us to rapidly analyse and report their experiences and concerns in a timely manner. Another limitation was that for some school responses were only provided by one participant so may not necessarily be representative of the whole school.

### Implications of findings

The findings of this survey provide educationalists and policy-makers with real-world frontline data to help make more informed decisions to ensure that educational settings remain open throughout the pandemic. Education staff, including teachers, are working hard to follow national recommendations to help keep schools safely open during the COVID-19 pandemic, despite most of them considering themselves to be at increased risk of COVID-19 because of their profession and being concerned for their own health. Parents too expressed concern about schools reopening and, while most were not worried about the health of their children, they were worried about their children transmitting the virus to others, including vulnerable household member. The impact of new COVID-19 variants with the potential for increased transmissibility will require careful monitoring when schools reopen during the Spring 2021 school term.^20^ As will the impact of mass testing using lateral flow devices that is currently being offered to staff and parents for school and home testing for SARS-CoV-2, a measure which may alleviate some of the concerns raised by parents staff and provide assurance.^21^

While most recommendations in the national guidance have been implemented to some extent in most schools, consistent concerns include difficulties in maintaining physical distancing within the school environment. This, along with difficulties of maintaining small bubble sizes following full reopening of schools, raises the question whether all staff and students should wear facemasks/covering in school,^22^ as has been implemented in other countries.^23^ Indeed, most parents appear to be supportive of children wearing masks in school.^1^ More generally, more studies are needed to assess the relative benefits of current infection control measures, including, but not restricted to, facemasks/coverings, so that future guidelines are more evidence-based.^23,24^

In particular, smaller class sizes, through blended in-school and home learning for example, would enable more effective implementation of the recommended infection control measures and provide additional reassurance for staff, parents and students.^25^ This would, however, only be possible if schools are provided with sufficient IT, computer hardware and internet support to allow the students to attend their classes online.^26^ We also identified a need to improve communications between policy makers and education staff in schools.^23^ Many staff members commented on some unrealistic recommendations in the national guidance, such as maintaining physical distancing and seating arrangements within the class, whilst attempting to bring all the children back to school in the current autumn term. Providing individualised and pragmatic support for schools that are unable to implement some specific measures, including financial support where needed, would help improve relationships and ensure optimal prevention practices in educational settings.^27^ By the same token, improved communication with parents, either directly by policy makers or through schools, would provide additional reassurance about the safety of their children attending school. Consistent messaging and using social media to reach younger people would also help communicate public health messages to promote behaviours that reduce COVID-19 transmission.^28,29^

Overall, there is growing evidence the risk of infections and outbreaks in educational settings correlate strongly with community SARS-CoV-2 infections rates in adults.^18,30,31^ Interventions to reduce local community infection rates, including local and national lockdowns without school closures as was recently implemented in England, not only reduced SARS-CoV-2 infections in adults but also in school-aged children. On-going surveillance in educational settings, however, remains critical due to/as a result of the changing landscape of the pandemic.

### Conclusion

Variable implementation of infection control measures and whilst the majority were not worried about returning to school, some parents and staff, were concerned about returning to school and the risk that posed to children, staff and household members. Following the closure of schools in January to March, the addition of regular mass testing to the suite of risk-reduction measures, some of these concerns may be alleviated. Continued monitoring of SARS-CoV-2 in schools, including the concerns for staff and parents, is required as society increasingly opens and the risks within schools alters.

## Data Availability

Data not available due to ethical/legal restrictions

## Data Availability

Data not available due to ethical/legal restrictions

## Acknowledgements

We would like to thank all the staff and parents who participated in the survey.

## Ethics approval

The sKID and sKIDsPLUS studies received approval from the PHE Research and Ethics Committee as part of PHE’s responsibility to investigate risk and transmission of SARS-CoV-2 among children in educational settings.

## Funding

None

## Conflicts of interest

None

## SUPPPLEMENT

**Supplementary Table 1:** Overall characteristics of schools participating in parent and staff surveys

|  | Parent |  | Staff |  |
| --- | --- | --- | --- | --- |
|  | n (%) |  | n (%) |  |
|  | Response | Non-response | Response | Non-response |
|  | N=56/152 | N=96/152 | N=76/152 | N=79/152 |
| **Type of school** |  |  |  |  |
| **Academy/free school** | 28 (51.9%) | 45 (49.5%) | 38 (54.3%) | 35 (46.7%) |
| **Community** | 15 (27.8%) | 28 (30.8%) | 21 (30.0%) | 22 (29.3%) |
| **Foundation/Voluntary** | 9 (16.7%) | 14 (15.4%) | 10 (14.3%) | 13 (17.3%) |
| **Independent** | 0 (0.0%) | 1 (1.1%) | 0 (0.0%) | 1 (1.3%) |
| **Special school** | 2 (3.7%) | 3 (3.3%) | 1 (1.4%) | 4 (5.3%) |
| **No of pupils on roll** |  |  |  |  |
| **<200** | 11 (19.6%) | 20 (21.3%) | 9 (12.5%) | 22 (28.2%) |
| **200-499** | 27 (48.2%) | 56 (59.6%) | 41 (56.9%) | 42 (53.8%) |
| **500-799** | 3 (5.4%) | 12 (12.8%) | 5 (6.9%) | 10 (12.8%) |
| **800-999** | 2 (3.6%) | 4 (4.3%) | 4 (5.6%) | 2 (2.6%) |
| **1000+** | 13 (23.2%) | 2 (2.1%) | 13 (18.1%) | 2 (2.6%) |
| **% of pupils on FSM** |  |  |  |  |
| **<5%** | 9 (16.1%) | 19 (20.2%) | 11 (15.3%) | 17 (21.8%) |
| **5-9%** | 8 (14.3%) | 6 (6.4%) | 8 (11.1%) | 6 (7.7%) |
| **10-19%** | 15 (26.8%) | 24 (25.5%) | 19 (26.4%) | 20 (25.6%) |
| **20-29%** | 16 (28.6%) | 19 (20.2%) | 19 (26.4%) | 16 (20.5%) |
| **30+%** | 8 (14.3%) | 26 (27.7%) | 15 (20.8%) | 19 (24.4%) |
| **% of persistent absence** |  |  |  |  |
| **<5%** | 14 (27.5%) | 19 (22.4%) | 18 (26.5%) | 15 (22.1%) |
| **5-9%** | 14 (27.5%) | 34 (40.0%) | 21 (30.9%) | 27 (39.7%) |
| **10+%** | 23 (45.1%) | 32 (37.6%) | 29 (42.6%) | 26 (38.2%) |

**Supplementary Table 2:**
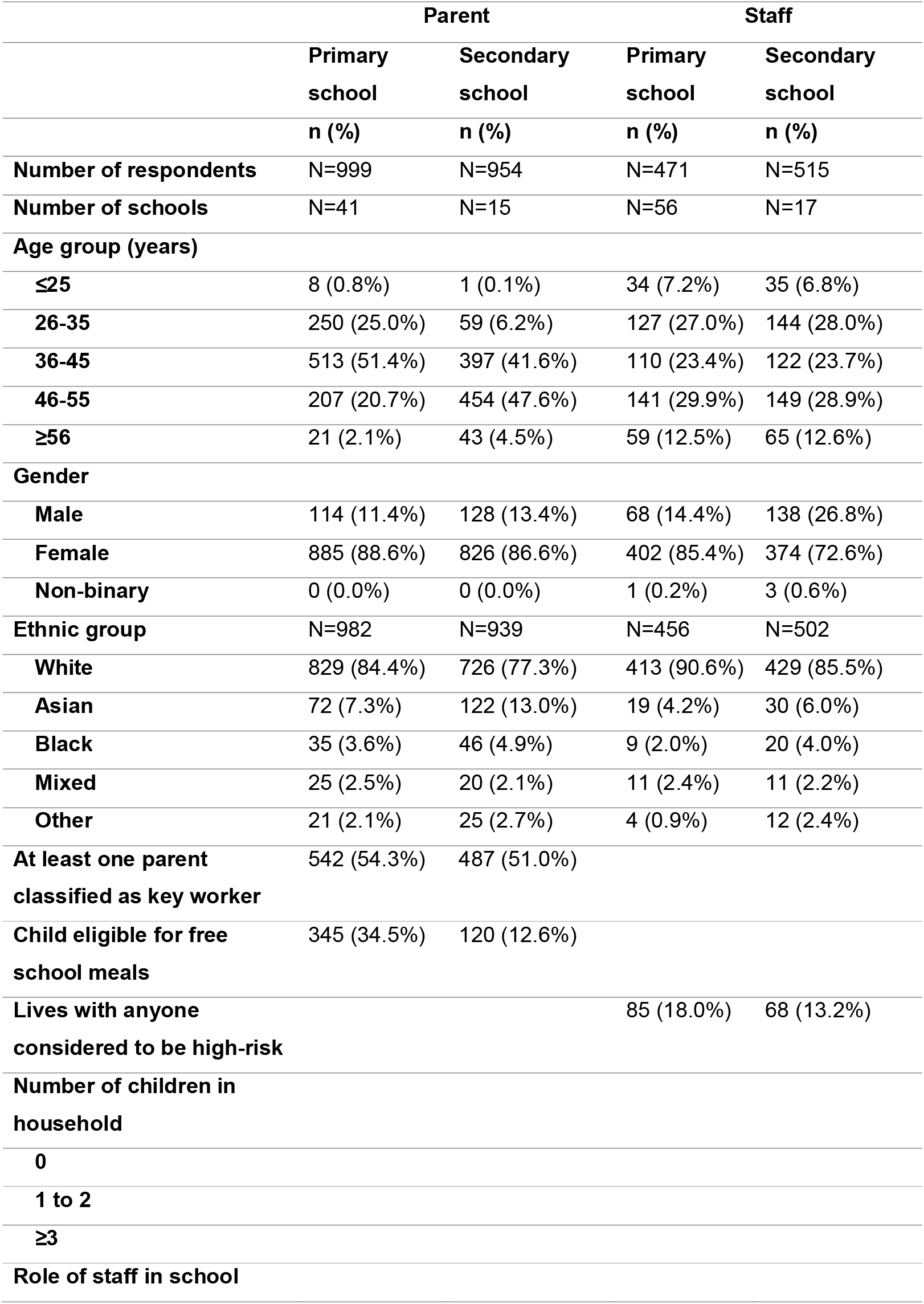
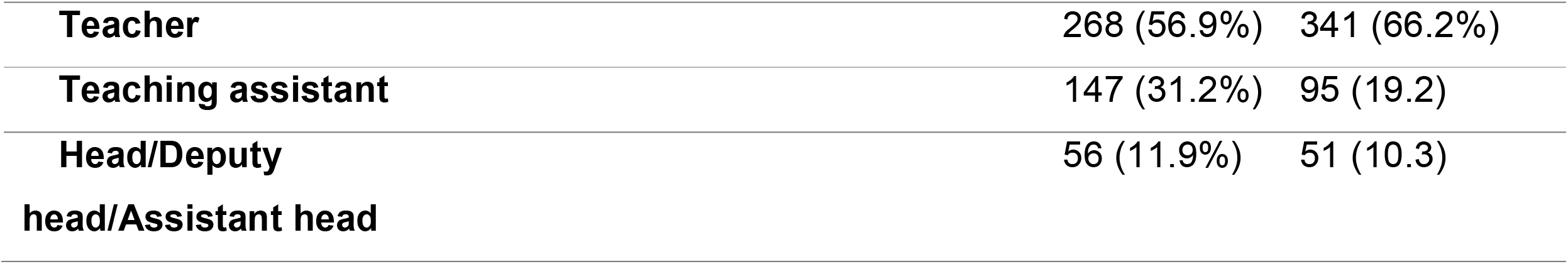
Parent and staff demographic and socioeconomic characteristics

## Notes

### Competing Interest Statement

The authors have declared no competing interest.

### Funding Statement

No external funding was received.

